# Temporal relationships between food insecurity, water insecurity, and generalized anxiety among rural Kenyans: seeking to clarify a climate-related syndemic

**DOI:** 10.1101/2023.11.10.23298356

**Authors:** Michael L. Goodman, Shreela Sharma, Dawit Woldu, Heidi McPherson, Ryan Ramphul, Stanley Gitari, Christine Gatwiri

## Abstract

**Background:** Increased water and food insecurity are one mechanism through which climate change can undermine global mental health. Understanding correlations between mental health and resource insecurity is imperative to support local adaptive responses to climate change.

**Objective:** We investigate temporal relationships between food insecurity, water insecurity, and generalized anxiety within rural Kenyans (n=152 adults) during a period of erratic rainfall.

**Method:** Using refined existing scales of food and water insecurity and generalized anxiety, we assess temporal relationships between these factors using cross-lagged panel analysis of survey data collected in October 2021 and October 2022 among participants in a community empowerment program.

**Results:** Food and water insecurity demonstrated significant, positive, reciprocal cross-lagged correlations. Generalized anxiety demonstrated significant, positive, reciprocal cross-lagged correlations with water insecurity. Food insecurity was not correlated with generalized anxiety within or between panels.

**Conclusions:** Supporting climate adaptation requires understanding temporal relationships between mental health and water and food security. Anxiety may reduce capacities to adapt to water insecurity, as well as be reduced by water insecurity. Food security may increase the capacity of households to adapt to water insecurity, though further research is required to establish causation and mechanisms for all observed temporal correlations in this study.

## INTRODUCTION

One mechanism through which climate change impacts the health of communities is through disruptions to rainfall and water security (Burke, Hsiang, & Miguel, 2015; Fjelde & Von Uexkull, 2012). Meta-analyses demonstrate altered precipitation patterns increase risk of interpersonal and intergroup conflict (Burke, Hsiang & Miguel, 2015). Erratic rainfall can impact crop productivity, potentially disrupting food systems in more climate-vulnerable communities (Mar et al., 2018; Simelton et al., 2011). An analysis of 2020 World Gallup Poll cross-sectional data found significant correlations between household-level food and water insecurity across Asia, Latin America, North Africa and sub-Saharan Africa, and concluded with a call for more research on pathways between food and water insecurity (Young et al., 2023).

Given the requirements of water for agricultural production, there is clear, potential causal directionality from water insecurity to food insecurity (Simelton et al., 2011; Mar et al., 2018). Smallholder farms, characterized by being two hectares or smaller in size, comprise 24% of agricultural area globally and contribute 30-34% to the total global food supply (Ricciardi, Ramankutty, Mehrabi, Jarvis & Chookolingo, 2018). The percentage of total agricultural land area comprised by smallholder farms is significantly higher in low- and middle-income countries (∼80%) compared to high-income countries - where about 50% of total agricultural land is comprised by smallholder farms (Lowder, Sanchez, & Bertini, 2021). As investments in irrigation and water maintenance is higher smallholder farms relative to total financial output, promoting resilience to fluctuations to rainfall disruptions is challenged across food systems served by predominantly by smallholder farms (Nakawuka, Langan, Schmitter, & Barron, 2018). In Kenya, for example, only 1.7% of smallholder farms have irrigation systems (Nakawuka et al., 2018). Accordingly, a reduction in rainfall by 50% in Meru County, Kenya contributed to a 70% reduction in maize production in 2016 (Nakawuka et al., 2018).

An evaluation of nationally-representative data across Kenya found 30% of households report spending over 75% of their household budget on food – with rural households spending 79% of their household income on food (vs. 62% average share of household income on food expenditure in urban settings; Mutea et al., 2022). Household caloric intake was lower than household caloric requirements in 40% of households – with substantially higher rates of caloric insufficiency in rural areas (Mutea et al., 2022). Across East Africa, rainfall variation predicts future rates of food insecurity according to International Phase Classification criteria (Coughlan de Perez et al., 2019).

Understanding how people cope with water insecurity can inform how food insecurity may contribute to water insecurity. Coping strategies for food insecurity, mirroring strategies used to cope with water insecurity, include limiting consumption, withdrawing from resource-sharing social obligations, and intensifying efforts to secure food – including increased labor, foraging, and selling assets (Wutich & Brewis, 2014). Illustrating linkages between food and water insecurity, study participants in western Kenya report they choose between purchasing food or water as a response to water insecurity (Collins et al., 2019). Better food security may reduce shocks introduced by climate factors, such as erratic and unpredictable rainfall, mitigating the experience of water insecurity.

Of the many health challenges subsequent to experiences water insecurity, recent research has found connections between water insecurity and poor mental health (Wutich, Brewis & Tsai, 2020). Theoretical basis for connections between water insecurity and worse mental health has been articulated through additional worry due to material deprivation, negative social evaluations, health costs; opportunity costs of fetching water; interpersonal conflict; and perceptions of unfairness (Wutich et al., 2020). A 2020 systematic review found food insecurity is correlated with higher depression, anxiety, and stress across ten different countries, though only one of these studies included longitudinal data (Pourmotabbed et al., 2020). A 2021 analysis of cross-sectional from 160 countries found food insecurity – defined as worry about household food adequacy, skipping meals, or facing chronic hunger – correlates with more symptoms of poor mental health and wellbeing (Elgar et al., 2021).

To promote clarity around how broader contextual and structural risk – such as erratic rainfall – contribute to mental health, researcher have begun promoting a syndemic view of water and food insecurity and mental health (Workman et al., 2021). Syndemic theory forwards that certain risk factors co-occur with sufficient frequency that these factors should be measured and analyzed in tandem as potential mediators of broader risk factors and health outcomes (Singer, 2000). Other syndemic conditions, typically considered as involving a biological status such as living with HIV, challenge mental well-being (Mendenhall & Singer, 2020). Emerging literature on effects of climate change and mental health implicate at least three types of climate-related pathways – through acute disruptions (e.g. hurricanes and wildfires), subacute or long-term changes (e.g. drought and heat stress), and the existential threat of enduring changes (e.g. rising sea levels, and potentially uninhabitable physical environments (Palinkas & Wong, 2020). While research on climate change and mental health may benefit from viewing other conditions affected by climate change with a syndemic perspective, we apply a syndemic approach within a socio-ecological framework to water insecurity, food insecurity and anxiety.

Socio-ecological frameworks consider the interactions of variables across multiple levels – individuals, nested within households, nested within larger communal, societal, environmental and temporal concentric circles (Kilanowski, 2017). In the case of the present study, we consider how data reflecting individual-level anxiety correlates with household food insecurity and water insecurity; water and food insecurity are impacted by variables at higher levels (e.g. rainfall and farming practice).

Disentangling temporal associations between food insecurity, water insecurity, and anxiety is necessary to inform preventive and treatment responses to the 301 million people living with anxiety disorders, the four billion people who face severe water scarcity during an average year, or the two billion people who face moderate or severe food insecurity globally (Aryal, Manchanda, & Sonobe, 2022; Mekonnen & Hoekstra, 2016; Boretti & Rosa, 2019; Yang et al., 2021).

Water and food insecurity have been associated with higher anxiety, though literature is conclusive about the nature and consistency of these associations (Brewis, Roba, Wutich, Manning, & Yousuf, 2021; Pourmotabbed et al., 2020). One earlier study shows seasonally-impacted food insecurity predicts higher anxiety (Hadley & Patil, 2008). Environmental stressors, and inadequate financial coping practices (e.g. food budgeting), exacerbate food insecurity among indigenous Inuit women (Beaumier & Ford, 2010). These two observations – that seasonal disruptions to food supply may animate anxiety, and coping practices improve resilience to food insecurity – suggest a potential reciprocal relationship between anxiety and food insecurity. General anxiety is characterized by higher presence of beliefs and mental schemas that may reduce the positive adaptability of individual behavior to larger threats like food or water insecurity (Koerner, Tallon, & Kusec, 2015; Mahoney et al., 2018). Pourmotabbed et al. (2020), however, found through a systematic review that food insecurity is inconsistently associated with water insecurity in cross-sectional studies. Linkages between water insecurity and anxiety have mostly been asserted through cross-sectional studies, though qualitative data suggest the need for longitudinal studies that advance the research field towards confirming or disconfirming causality and related mechanisms (Wutich, Brewis, & Tsai, 2020).

One potential source of confounding between food insecurity, water insecurity and anxiety is worry is considered part of the definitions of most measures of food and water insecurity, and a key feature of anxiety disorders (Ashby, Kleve, McKechnie & Palermo, 2016; Young et al., 2021; Mathews, 1990). Clarifying temporal associations between food insecurity, water insecurity, and anxiety likely requires controlling for generalized anxiety within panel data where food or water insecurity measures include worry. Clarifying this temporal order can support etiological understandings of environmental impacts to anxiety, help tailor interventions, and provide testable models to predict population-level mental health impacts of identifiable, population-level predictors such as threats to food and water security. Understanding the intersections between food insecurity, water insecurity, anxiety and linkages to environmental factors, may provide insights to improving overall prosperity and wellbeing (Baldwin-Cantello et al., 2023).

### Study Aim

This study seeks to clarify temporal ordering between anxiety, food insecurity and water insecurity among rural Kenyans in Meru County between October 2021 and October 2022.

## METHODS

### Study Location

Figure 1a shows the location of Meru County within Kenya. This study is located in North and Central Igembe sub-counties of Meru County which is located just north of Mt. Kenya, in the center of the country. Figures 1b-e show rainfall variability and food security during the study period. As shown in Figures 1b and 1c, the seasonal accumulation of rainfall between March to December 2021 as a percentage of normal precipitation fluctuated greatly across East Africa. The study location transitioned from receiving 100% of expected rainfall between March-December 2021 to receiving 60% of expected rainfall between March and December 2022 (United States Department of Agriculture, 2023). Figures 1d and 1e show food insecurity in October 2021 across Kenya, categorized by sub-county. In October 2021, food security North and Central Igembe sub-counties was considered “stressed” and in October 2022, food security in these sub-counties was considered “in crisis” (Famine Early Warning Systems Network, 2023).

**Figures 1a-e.**
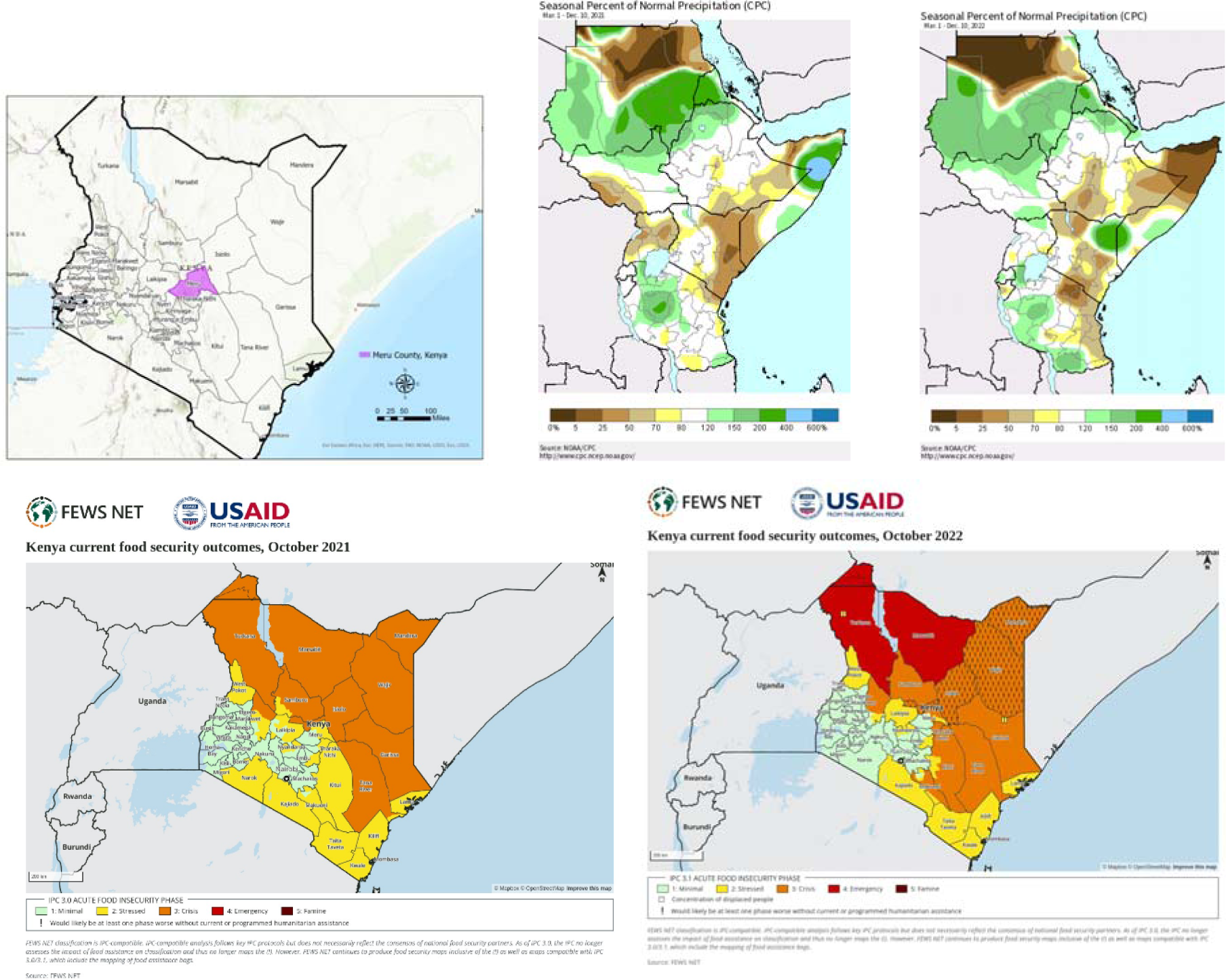

### Study Setting

Data for this study were collected from participants in a community-based empowerment program, previously showing efficacy to improve mental health, parenting behaviors, income, and reintegration outcomes for street-involved children and youth (Goodman et al., 2021a; Goodman et al., 2023a; Goodman et al., 2023b). The interventional design combines group-based microfinance practices, leadership development, and a novel behavioral health curriculum to animate values-based growth (Goodman et al., 2023b). An index of lending group-affiliated interpersonal trust, norms of reciprocity, sense of belonging, and cohesion previously demonstrated cross-sectional associations with lower food and water insecurity (Goodman, Elliott, Melby & Gitari, 2022). Survey-based questionnaire data were collected in October 2021 (T1) and October 2022 (T2).

### Program recruitment

Program participants were recruited to the intervention through a combination of active and passive recruitment. Active recruitment begins at the village-level through identification of families of children living on the streets or families engaged in HIV care at Ministry of Health clinics (Goodman et al., 2023b). These actively recruited, index families within villages recruit 25-29 other families to begin a microfinance group. Additional families join the program through passive recruitment, driven mostly by word of mouth (Goodman et al., 2021b). Participants are thus nested within groups, who are nested within villages. There are 10 villages represented in the current study.

### Participant selection

After groups are established, program evaluation is conducted to ascertain trends in key variables across the duration of active program support. Participants in the current study were randomly selected during routine group-based microfinance activities using a random number selection process. All willing participants (98.4% of all participants) are invited to draw a folded piece of paper with “1” (selected) or “0” (not selected) from an opaque bag. Each established group within a village is invited to participate in the evaluation process. There are 152 individual participants with paired T1 and T2 data in the current study.

### Study Design

This data follows an interventional cohort design, with two waves of collected data.

### Measures

The interviewer-delivered survey questionnaire was developed in English, translated into the local language (Kimeru), and back translated to English for comparison and refinement. The survey questionnaire was administered by a team of trained local experts affiliated with two Kenyan universities within the study county.

### Primary measures

This study analyzes cross-lagged relationships between three primary variables – generalized anxiety, water insecurity and food insecurity. Responses to all primary variables are continuous averages of item-level responses.

### Generalized anxiety

Generalized anxiety was measured using the GAD-7 reported on a 4-point Likert-type response reflecting a range of “not at all” to “nearly every day” experience of symptoms of generalized anxiety (α=0.82 at T1; α=0.82 at T2; Spitzer, Kroenke, Williams, & Lowe, 2006). The GAD-7 is a 7-item measure using a 2-week recall period, prompting respondents to report the frequency of experiencing nervousness, worry, difficulty relaxing, and irritability. The GAD-7 has previously been validated in Kenya and other global populations (Goodman et al., 2022; Dhira, Rahman, Sarker, & Mehareen, 2021).

### Water insecurity

Household water insecurity was measured using the first factor (17-items) of the HHWI scale; responses to items on the HHWI are reported on a 5-point Likert-type format reflecting household water insecurity ranging from “never” to “always” (α=0.95 at T1; α=0.95 at T2; Boateng et al., 2017). The HHWI was developed in western Kenya, which motivated its selection in the current study. The 17-items cover worry, time devoted to fetching and related opportunity costs, insufficiency of water for household- and self-care, social avoidance due to feeling too dirty, conflicts with neighbors due to limited water, experiencing thirst, and changing diet to accommodate for low water access during the previous 4-weeks. Through principal components factor analysis, the item with the lowest factor loading (0.44) was “In the last 4 weeks, how frequently have you worried about the safety of the person getting water for your household?”; the item with the highest factor loading (0.84) was “In the last 4 weeks, how frequently have you or anyone in your household had to go without washing their body because there wasn’t enough water?”

### Food insecurity

Food insecurity was measured using an item reduction method of the 9-item HFIAS, which has previously been validated in multiple sub-Saharan African countries (Musemwa et al., 2015; Otekunrin et al., 2021). Following Hager et al. (2010), we assessed items within a previous HFIAS sample (α=0.91; Goodman et al., 2022) with the strongest factor loadings among items covering (1) frequency changing diet due to insufficient food and (2) frequency going hungry due to insufficient food. The HFIAS uses a 4-week recall compared to the 12-month recall informing the Hager et al. (2010) approach, contributing to greater responsiveness to near-term fluctuations in the food environment. In previous data, this 2-item method had higher correlation with the full scale (r=0.93) than the previously cross-culturally validated 3-item scale reduction of the HFIAS (r=0.89; Deitchler, Ballard, Swindale, & Coates, 2010). As such, we utilized a 2-item scale of food insecurity reflecting alterations in consumption and experience of hunger within the past 4 weeks (r = 0.55 at T1 and r=0.61 at T2). The item response format for food insecurity is a 4-point Likert-type response reflecting “never” to “often.”

### Control variables

To address potential confounding from unobserved correlations between primary variables and sociodemographic factors, we assessed T1 measures of wealth, income, formal education, age, and gender. Wealth was a composite index of 12 common household items (KR20 = 0.71). Wealth indices are a valid method of ascertaining one dimension of socio-economic position, though distinct from income (Poirier, Grepin, & Grignon, 2020). Household income was measured as a continuous estimate of monthly income in Kenyan shillings. Formal education was measured and analyzed as a continuous number reflecting number of years of formal school attendance. Age was recorded as a continuous measure of years since birth. Gender was considered a binary variable – man or woman.

### Statistical analysis

We describe all variables included in the analysis, indicating mean and standard deviations. We compare equivalence of primary measures between T1 and T2 using Student t-tests.

We calculated correlation matrix of all primary and control variables using Spearman’s rank correlation coefficient.

Primary analyses were conducted using a cross-lagged panel analysis calculated using Structural Equation Modelling (SEM) with all primary measures at T1 and T2, adjusting for T1 control variables. All possible pathways were included between T1 and T2 primary measurements, controlling for correlation between all primary variables within T1 and T2 and assessing for control measures at T1. All pathways with p<0.2 were excluded, and the model recalculated until all correlations were significant at p<0.1. We only interpret correlations at p<0.05 as statistically significant and meriting discussion.

All statistical analyses were conducted using STATA v.16.1 (StataCorp, 2019).

### Ethical considerations

All study objectives and protection of human subjects protocols were reviewed and approved by the Institutional Review Boards at the Institutional Review Boards at the Kenya Methodist University and the University of Texas Medical Branch in Galveston, TX. All participants provided informed written consent that emphasized the voluntary and confidential nature of the study prior to participating in the study. Compensation ($1) was given to participants’ microfinance group on behalf of each study participants, following a practice designed by community members interested in ensuring equitable distribution of available resources following a random selection.

## RESULTS

Table 1 shows the univariate descriptions of primary variables at T1 and T2, and control variables at T1. Food insecurity remained unchanged between T1 and T2, with mean close to 1.4 at each wave. Water insecurity significantly declined between T1 and T2, decreasing from mean 1.4 (sd: 0.9) to 1.3 (sd: 0.9) on between T1 and T2. Anxiety remained without significant change between T1 and T2 (mean: 1.1).

**Table 1:**
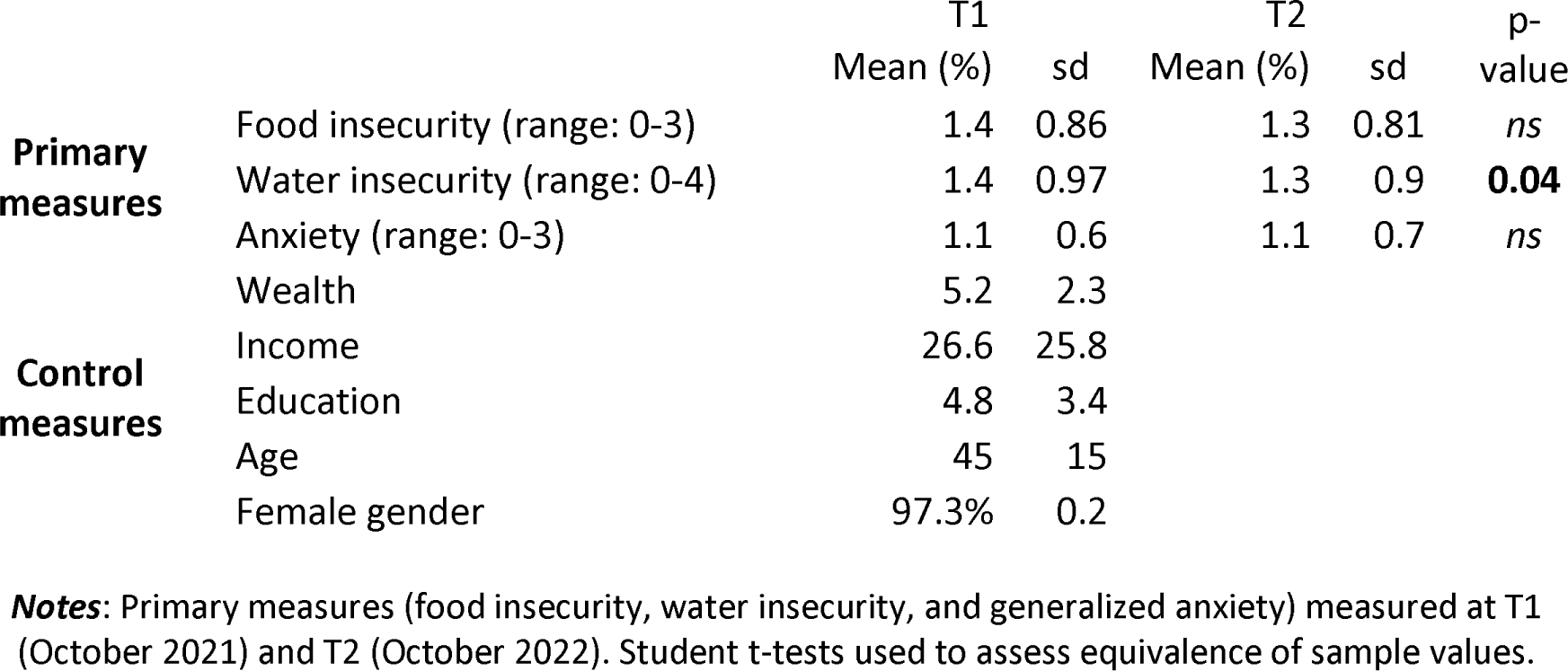
Univariate characteristics and comparisons of primary values between T1 and T2 (n=152)

Mean items owned on the household wealth index was 5.2 (sd: 2.3) at T1. Mean income was 3033 KSH per month. Mean age was 45 years (sd: 15) at T1. Over 97% of respondents were women.

Table 2 shows the correlation matrix of all assessed, continuous variables. As shown, food insecurity (T1) was significantly correlated with food insecurity (T2; r=0.41, p<0.001), water insecurity (T1; r=0.35, p<0.001), water insecurity (T2; r=0.3; p<0.001), wealth index (T1; r=-0.3; p<0.001), age (T1; r=0.3, p<0.001). Food insecurity (T2) was significantly correlated with water insecurity (T1; r=0.21, p<0.01; T2; r=0.36, p<0.001), wealth (T1; r=-0.35, p<0.001), income (T1, r=-0.27, p<0.001), age (T1; r=0.23, p<0.01), and years of education (T1; r=-0.16, p<0.01). Water insecurity (T1) was significantly correlated with water insecurity (T2; r=0.49, p<0.001), and generalized anxiety (T2; r=0.25, p<0.001).

**Table 2.**
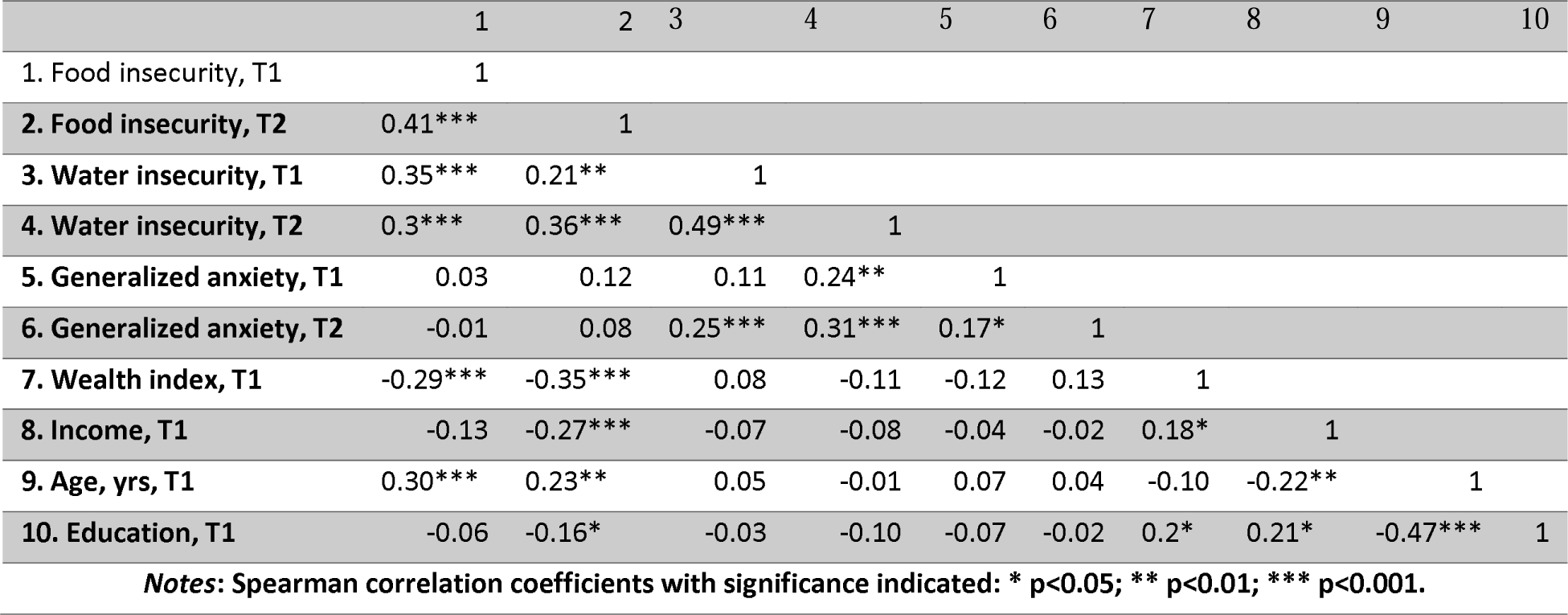
Spearman correlation coefficient matrix.

Figure 2 displays the cross-lagged panel analysis. Worse food insecurity (T1) predicted worse water security (T2; r=0.19, p<0.01) and food insecurity (T2; r=0.26, p<0.001). Worse water insecurity (T1) trended towards significantly higher food security (T2; r=0.13, p=0.1), worse water insecurity (T2; r=0.41, p<0.001), and more generalized anxiety (T2; r=0.19, p<0.05). Worse generalized anxiety (T1) predicted worse water insecurity (T2; r=0.17, p<0.01), and worse subsequent generalized anxiety (T2; r=0.15, p<0.05).

**Figure 2:**
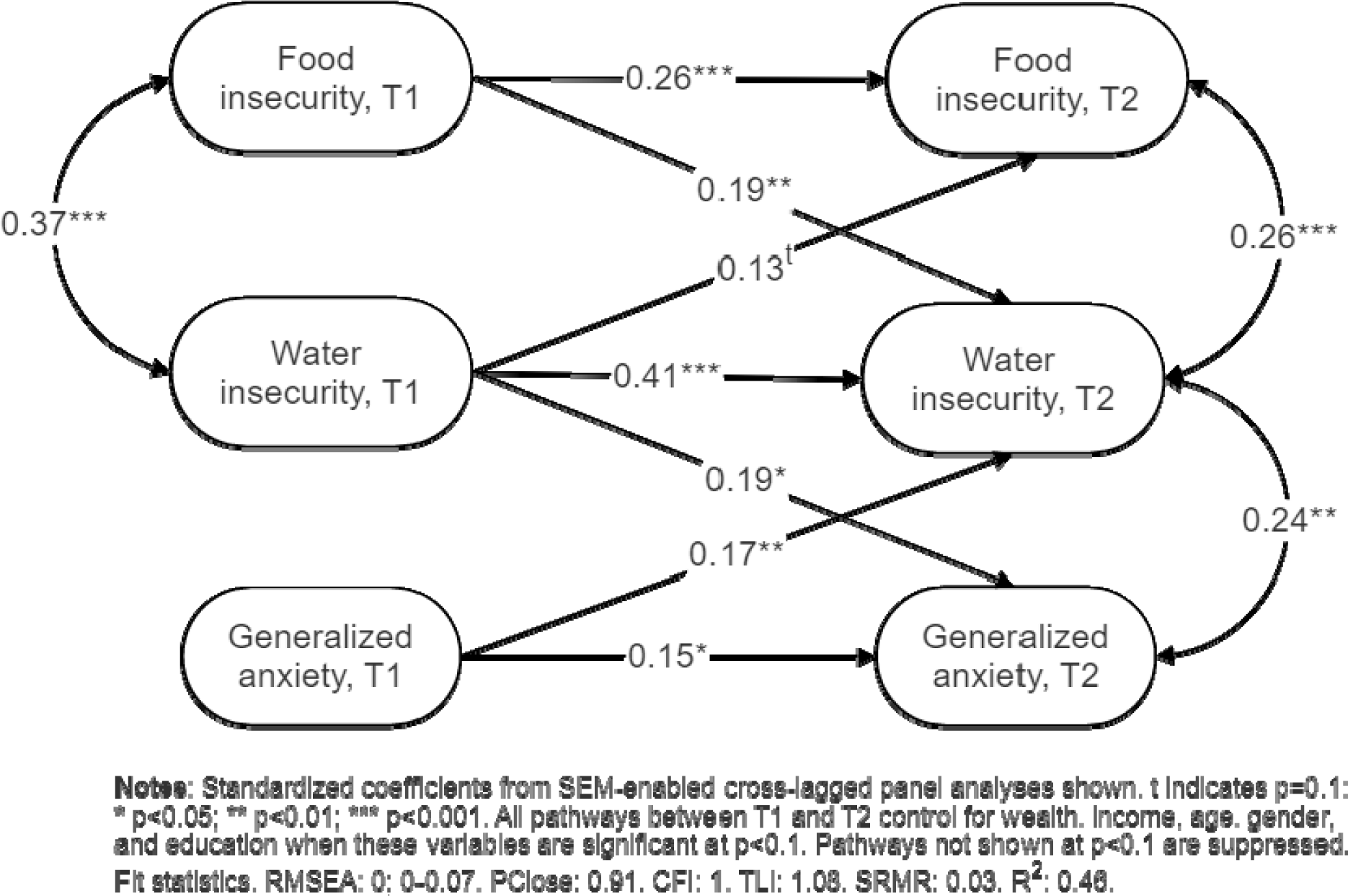
Cross-lagged panel analysis using SEM with standardized coefficients.

## DISCUSSION

This study sought to clarify temporal relationships between food insecurity, water insecurity, and generalized anxiety within an interventional cohort in rural Meru County, Kenya. We found reciprocal bivariate correlations between water and food insecurity at T1 and T2. Controlling for within-panel correlations at T1 and T2 demonstrated food insecurity (T1) was more strongly predictive of water insecurity (T2) than water insecurity (T1) predicted food insecurity (T2). These findings suggest baseline better food security may reduce the impact of future water instability – including, potentially, the impact of irregular rainfall. The mechanisms underlying such a protective benefit require more investigation, though existing research indicates one way people cope with water insecurity is through decreasing consumption of other resources (Ventkataramanan et al., 2020); higher baseline food security may indicate access to resources that can be reduced in times of higher water insecurity.

Beyond supporting the syndemic resource insecurity hypothesis, this study indicates the need for future research to explore potential overlapping socio-biological complications related to syndemic resource insecurity. As water insecurity was correlated with food insecurity within waves, and was correlated with later food insecurity at near significance, the food-water insecurity syndemic may impact physical health by motivating families to engage in unsafe water consumption potentially leading to infection, and through inadequate calories, macronutrient or micronutrient consumption (Venkataramanan et al., 2020; Misselhorn & Hendricks, 2017). A cross-sectional study in 16 low- and middle-income countries found water insecurity is perceived to directly affect infant breastfeeding and non-breastmilk feeding through multiple pathways (Schuster et al., 2020).

Findings suggest generalized anxiety may reduce the capacity to adapt to future disruptions to water security. It would be reasonable to consider that people with generalized anxiety have a higher propensity to worry about water security, which could be reflected in the water insecurity measure. However, controlling for within-panel correlations between anxiety and water insecurity reduced this threat to internal validity. The role of anxiety to support adaptive coping requires more research within the study context – and similar contexts across sub-Saharan Africa and other areas sensitive to disrupted rainfall and other sources of water insecurity.

Future research should explore how water insecurity may impact beliefs and mental schemas that reduce positive adaptation to future threats that characterize the experience of anxiety (Koerner et al., 2015). If the observed cross-lagged panel analyses reflect true population-level dynamics, climate disruptions to water security may engender or exacerbate vicious cycles between climate, behavioral and cognitive adaptation, and generalized anxiety. Understanding the dynamics between social and climate related development efforts, is important to ensure successful synergies that promote water and food security.(United Nations Department of Economic and Social Affairs (UNDESA), United Nations Framework Convention on Climate Change Secretariat (UNFCCC), 2023) Even climate-supportive actions, such as shifting to renewable hydropower, can impact water ecology, water insecurity, and food insecurity, without adequate community engagement and forethought (Baldwin-Cantello et al., 2023; United Nations Department of Economic and Social Affairs (UNDESA), United Nations Framework Convention on Climate Change Secretariat, 2023). Given the estimated increase in number of people who will face water insecurity globally from 4 billion to 6 billion people by 2050 (Boretti & Rosa, 2019), understanding reciprocal dynamics between water insecurity and behavioral and cognitive coping mechanisms is essential.

We were surprised food insecurity was not associated with generalized anxiety within or between time points – in neither SEM-controlled nor bivariate correlations. While these observed null findings are congruent with the systematic review reported in Pourmotabbed et al. (2020), we suggest caution in over-extrapolating from these observations. The food insecurity measure used in this study, chosen to reduce response burden on participants in an active programmatic intervention, did not include worry about food access. It is possible that beyond controlled, within-panel correlations between anxiety and food insecurity, other correlations between generalized anxiety and food-specific worry exist. These correlations would not be identified within the present data, as the food insecurity measure excluded food-specific worry. It is possible that generalized anxiety does not contribute to, or emanate from, food insecurity – though given the high global prevalence of anxiety disorders and food insecurity, such conclusions require more confirming support. Finally, it is possible that there are conditions in which food insecurity and anxiety are associated, and conditions in which they are not. This remains a topic open within the literature.

There have been few longitudinal studies investigating generalized anxiety, food insecurity and water insecurity. These findings suggest dynamic interactions between food and water insecurity, and generalized anxiety and water insecurity. These dynamics should be further explored within the context of seasonal variations in water and food security, and explore whether and how mental health conditions impact climate adaptation, syndemic water and food insecurity mutually interact over time, environmental conditions exacerbate mental health, and interventional mechanisms to invert potentially antagonistic relationships between these conditions at population-levels.

### Limitations

These data were collected from an interventional cohort with research priorities beyond water and food insecurity – namely, the identification of community- and individual-level mechanisms to promote adaptation that include elements of communal psychology, positive psychology, and psychological flexibility. This broad interest can produce a rich dataset, and support novel identification of potentially promotive associations within an under-studied population, the breadth can prevent answers to more specific questions – such as observed antagonistic relationships between anxiety and water insecurity, and between food and water insecurity. Future research should explore specific coping mechanisms used by people in water and food insecure environments globally, and the psychological determinants and impacts of these coping behaviors.

Self-reported data are known to be susceptible to certain biases such as recall and social desirability biases. Recall bias could have been introduced through asking respondents to report on water and food insecurity during the previous 4 weeks, and could be mitigated through daily journals. The use of external, objective measures – such as rainfall data and the presence and distance of non-surface water sources – could further reduce recall bias. Social desirability bias may have, within panels, introduced a deflation of resource insecurity, as resource insecurity is associated with shame due to experience of social failure or resorting to socially unacceptable methods to secure resources (Coates, Swindale, & Bilinksy, 2007). Social desirability bias would have to differentially affect how respondents with higher need for social acceptance report between T1 and T2 to impact the primary correlations of concern to this study - those that appear between survey waves. Future studies should establish village-randomized or propensity-matched controls to observe temporal dynamics between variable food and water security, and anxiety.

There is also an opportunity to better understand the role of stigma and its impact on anxiety in relation to food and water insecurities. Stigma is root cause factor in population inequities, and has been found to play a role exacerbating existing inequities both at the structural and individual levels.(Earnshaw & Karpyn, 2021)

Furthermore, we acknowledge climate change and food-related intercommunal conflict could influence anxiety. Recent studies from the Sahel region west Africa indicated harsh climate conditions could lead to intercommunal armed conflict by acting as “threat multiplier” (Schon, Koehnlein and Koren, 2023; Koren, Bagozzi and Benson 2021). Schon, Koehnlein and Koren (2023) argue regions where there is seasonal climatic fluctuation are at greater risk of armed or intercommunal conflict, because of communities’ lack of adaptation, than communities with permanent harsh climate. The site of this study in Kenya falls within this erratic seasonal climate where food insecurity occurs quite frequent (Perez et al. 2019; Mutea et al.2019). Future study should explore how communal conflict related to climate change and food insecurity impacts anxiety.

## CONCLUSIONS

There are currently 301 million people living with an anxiety disorder, 4 billion people facing severe water scarcity at some point throughout the year, and 2 billion people facing moderate or severe food insecurity globally. Each of these estimates is expected to increase, particularly driven by the near- and long-term effects of climate change. Climate adaptation requires understanding temporal relationships between these syndemic elements, which we aim to support through cross-lagged panel analyses of interventional cohort data collected from rural Meru County, Kenya (n=152 adult participants). Findings suggest potential antagonistic relationships between food and water insecurity, and generalized anxiety and water insecurity. Future research should utilize qualitative methods to understand how people experience water and food insecurity, particularly how prior food security may help mitigate later water insecurity. Also, psychological, social, and behavioral determinants of adaptability to climate disruptions is required. Finally, future research should help link objective metrics of rainfall disruptions, and the food environment to self-reported responses.

## Data Availability

All data produced in the present study are available upon reasonable request to the authors

